# Carpal tunnel syndrome with shoulder symptoms; is it a different entity? A prospective cohort study comparing recovery patterns following decompression

**DOI:** 10.1101/2023.01.24.23284937

**Authors:** G N S Ekanayake, S R Manilgama

## Abstract

**Introduction:** Carpal tunnel syndrome (CTS) may present with atypical proximal symptoms other than hand symptoms. Among proximal symptoms, shoulder symptoms are most distant to the site of compression and leads to a battery of investigations to exclude other pathologies. The recovery patterns of proximal symptoms have not been studied following carpal tunnel decompression.

**Method:** A prospective cohort study was conducted to compare the recovery patterns between subgroup-A; CTS patients (n=55) with shoulder symptoms, with subgroup-B; patients (n=55) without shoulder symptoms. A pre-tested questionnaire was administered prior and after the surgery and information was gathered on day-7, day-14, and day-21.

**Results:** Of 110 patients, 81.8% were females. The mean age(±SD) of the total population was 49.2(±10.6), that of subgroup-A was 48.7(±10.3), and subgroup-B was 49.7(±11) years. They were predominantly right-handed (82.7%), 73% had their dominant hand affected while 74.5% were affected bilaterally at presentation. All had hand symptoms: numbness in 94.5%, pain in 85.4%, tingling of hand in 66.3% and all-three symptoms in 57.2%. In subgroup-A, pain was the predominant shoulder symptom which appeared before hand symptoms in 34.5%, after in 47.3% and simultaneously in 18.2%. It was aching-type in 91%. The pain radiated proximally (neck and scapula) in 61.8% and to the arm in 81.8%.

The recovery rates (RR) of hand symptoms following decompression on day-7 were 41.8% in subgroup-A and 40.0% in subgroup-B. RR of hand symptoms in subgroup-A and subgroup-B on day-14 were 72.7% and 70.9%, while on day-21 were 83.6% and 85.5% respectively. RR of shoulder symptoms in subgroup-A were 38.2% on day-7, 65.4% on day-14, and 78.2% on day-21.

**Conclusion:** Near identical RR of hand and shoulder symptoms were seen between the two subgroups after decompression, which indicates CTS with shoulder symptoms is not a different entity.

## Introduction

In the early twentieth century, carpal tunnel syndrome (CTS) was recognized as acroparesthesia. In 1906 Farquhar Buzzard and W. W. Keene suggested treating this with resecting the first rib (1). This theory of brachial plexus plexopathy persisted till the mid twentieth century. Subsequently, several surgeons started publishing their results to show a more distal reason for compression (2). This treatment focused on the carpal tunnel as the point of compression. A landmark paper by Cannon and Love in 1946 described the surgical treatment at the wrist level for CTS (3). Later, numerous studies by Brain and Phalen popularized the current understanding (4). The term “carpal tunnel syndrome” first appeared on print in 1953 by Kremmer et al (5).

Atypical proximal symptoms may have led to the uncertainty of the site of compression in CTS. Among atypical proximal symptoms, shoulder symptoms are the most distant to the site of compression and clinically more relevant (6). The shoulder symptoms in patients with CTS have been reported to be around 6.3 to 19% in different populations (7,8). The presence of atypical proximal symptoms and the different characteristics of shoulder symptoms are thought to be due to different pathophysiological reasons (9). The median nerve has three main anatomical variations at its formation, different patterns of branching and variable cross communications with other nerves (9, 10).

The specific treatment for CTS is decompression of the carpal tunnel at the wrist. Though the recovery patterns of distal hand symptoms are described following decompression, the data on recovery from proximal symptoms are scarce. This study mapped the recovery patterns seen among CTS patients with shoulder symptoms in comparison to those without shoulder symptoms. We also studied the characteristics of proximal symptoms including shoulder symptoms. Study of the recovery patterns of proximal symptoms following decompression helps to indirectly prove the causal relationship of origin of symptoms and potentially minimise unwanted and expensive investigations done to exclude other pathologies when a patient presents with shoulder symptoms due to CTS.

## Method

This prospective cohort study was conducted from January 2018 to January 2019 at the department of plastic surgery of a tertiary care institution in Sri Lanka. Patients diagnosed with CTS who would undergo decompression surgery were recruited for the study. They were referred from the departments of medicine and neurology after evaluation for CTS. Each one of them had undergone electromyogram (EMG) and nerve conduction study (NCS) to confirm CTS and to exclude other neuropathies. An individual who had clinical syndrome of median nerve (MN) compression confirmed by neuro-electrophysiological study was considered as a case of CTS. Comprehensive neck and upper limb examination was carried out by a trained physician. Cervical X-ray and ultrasound examination of the shoulder joint was carried out when clinically indicated. The following categories were excluded from the study: patients with diagnosed cervical radiculopathy, two or more compression neuropathies, other painful pathologies e.g., trigger finger, post trauma or active synovitis, active inflammatory arthropathies. All the subjects who fulfilled the above criteria and consented for the decompression surgery were enrolled. Informed written consent was taken for the study and consent for surgery was taken separately. A pre-tested, interviewer administered questionnaire was designed to extract necessary data from the individual subject prior to the surgery. The latter part of the questionnaire was completed following the decompression surgery over the phone. Symptoms of CTS were categorised according to the two regions involved: hand/forearm symptoms (below elbow) or shoulder/arm symptoms (above elbow). Patients were divided into two groups accordingly: both shoulder/arm and hand/forearm symptoms (subgroup-A) and patients with only hand/forearm symptoms (subgroup-B). Symptoms were further studied in detail in relation to onset, radiation, severity, nature, resolution, and associated other symptoms.

Post-op evaluation was done on day-7, day-14, and day-21. The resolution of symptoms following the surgery were recorded. Null-hypothesis of “there was no difference between the two groups in the recovery pattern after decompression surgery” was tested. Statistical Package for Social Sciences (SPSS) software version 20 was used for the analysis of data. Descriptive statistics were used to describe population parameters. Complete recovery rates in the subgroups at the three points were calculated. Significant differences between subgroups were calculated using the chi-square test. The ethical clearance was obtained from the ethical review committee of the institution.

## Results

Out of 110 patients, 81% were females (female: male ratio was 4:1). The mean age (±SD) was 49.2 (±11) years. They were predominantly right-handed (83%). Carpal tunnel syndrome was evident bilaterally at presentation in 83% while unilateral in 17%. The condition had predominantly affected the dominant hand in 73%. The population was divided into subgroup-A (n=55) and subgroup-B (n=55). The mean (±SD) age of subgroup-A and subgroup-B were 48.7 (±10.3) years and 49.7 (±11) years respectively. The prevalence of diabetes mellitus among subgroup-A was 18.2% while subgroup-B was 16.4%. Hypertension was reported as 18.2%, while hypothyroidism was reported as 3.6% in each subgroup.

### Hand symptoms

Hand symptoms were present in all the patients. The reported symptoms were numbness, pain and tingling sensation of the hand which were reported in 95%, 85.4% and 66.3% respectively. All three symptoms were present in 57.3%.

### Shoulder/arm symptoms

Patients with both shoulder and hand symptoms were identified as (n=55) subgroup A. Pain was the only symptom related to the shoulder region among them. Shoulder pain appeared before, after, and almost simultaneously with hand symptoms in 34.5%, 47.3% and 18.2% respectively. Shoulder pain extended further proximally to the neck and/or scapular region in 14.5%. The majority (81.8%) experienced arm pain in addition to shoulder pain. Shoulder pain was mild in 12.7%, moderate in 52.7% and severe in 34.5%. The majority (91%) experienced an aching type pain while the rest had pricking type (5.4%) or burning type (3.6%).

### Decompression outcome

Recovery rates of hand symptoms were recorded independently from recovery rates of shoulder symptoms on day-7, day-14 and day-21 following carpal tunnel decompression. Then the recovery rates of hand symptoms were compared with shoulder symptoms in subgroup A. Recovery rates of distal and proximal symptoms were nearly identical and not statistically significant (table 1).

**Table 1:** Complete recovery rates of hand and shoulder symptoms of patients with both symptoms in subgroup A (n=55)

|  | Day 7 | Day 14 | Day 21 |
| --- | --- | --- | --- |
| Hand symptoms recovery rate (%) | 41.8 | 70.9 | 85.5 |
| Shoulder symptoms recovery rate (%) | 38.2 | 65.4 | 78.2 |
| p value | 0.69 | 0.54 | 0.32 |

Recovery rates of hand symptoms in day-7, day-14 and day-21 following carpal tunnel decompression were recorded and compared between subgroup A and subgroup B. Recovery rates were not significantly different between the two subgroups in our study (table 2).

**Table 2:** Complete recovery rates of hand symptoms in the two subgroups (Subgroup A - patients with both shoulder and hand symptoms, subgroup B - patients with only hand symptoms)

|  | Day 7 | Day 14 | Day 21 |
| --- | --- | --- | --- |
| Subgroup A - hand symptoms recovery rate (%) n=55 | 41.8 | 70.9 | 85.5 |
| Subgroup B - hand symptoms recovery rate (%) n=55 | 40.0 | 72.7 | 89.1 |
| p value | 0.84 | 0.83 | 0.56 |

## Discussion

Our study showed near identical complete recovery rates of hand symptoms in CTS patients with or without shoulder/arm symptoms following decompression surgery. Recovery rate of hand symptoms on day-7 among patients without shoulder symptoms was 40.0% whilst among patients with shoulder symptoms was 41.8%. Recovery rates of hand symptoms were 72.7% and 70.9% on day-14, while recovery rates were 89.1% and 85.5% on Day-21 respectively. Complete recovery rates of shoulder symptoms on day-7, day-14 and day-21 were 38.2%, 65.4%,78.2% respectively.

A wide variation in clinical presentation of CTS may have contributed to the historical confusion in describing the syndrome. Clinical symptoms of the hand remain consistent and well understood. In our study population, the presence of hand symptoms was a universal finding among participants. In previous studies, paresthesia/numbness of hand was reported in >95% of patients with CTS (11,12,13). Similarly in our study, symptoms of numbness, pain and tingling of hand were reported in 95%, 85.4% and 66.3% respectively.

However, the symptoms of CTS are not only limited to the hand. These compression symptoms may extend proximally up to the shoulder girdle. Out of proximal symptoms the shoulder symptoms remain poorly described. The frequency of shoulder symptoms varies from one study to another from 6.3-19% in different populations. (7)

Patients belonging to subgroup A had symptoms not only limited to the hand region but extending proximally to the shoulder/arm region. All of them had pain in the shoulder, but the nature, severity and the timing were different. There are limited reports on the character of the shoulder symptoms up to now. This group of patients have described it as an aching type of pain. It is quite different from the hand symptoms. At the same time the majority has described it as a moderate to severe type of pain. Character of radiation is poorly understood, and the timing of the symptom may vary. In our population, shoulder pain appeared in one third of patients (34.5%) before hand symptoms. Rest of the population reported shoulder symptoms after and almost simultaneously with hand symptoms. They described the pain as radiating towards the arm, neck or even scapula. In 1973, Kummel et. al. described the presence of radiating shoulder pain in CTS (8). The presence of shoulder pain could alert the clinician to an alternative diagnosis or additional pathology. Therefore, proximal symptoms can take the patient and the clinician towards a battery of expensive, unnecessary and inconclusive investigations.

This referred pain or recently described retrograde neuropathic pain (RNP) is thought to have a neurological reason. There are mainly three theories explaining the RNP. The convergence theory is a phenomenon that has been demonstrated in various other nerves (14). It explains the possibility of combining specific pathways that carry sensory information from related regions to the brain. When extrapolating this to the MN, the sensory pathway of the hand and shoulder could be carried in the same nerve (15). Evidence from cadaveric studies for this phenomenon is limited. However, it could be explained by the Hiltons law applied to the musculocutaneous nerve (MCN). The mystery of a subset of patients with shoulder pain could also be explained by the existence of interconnections between MCN and MN as described in cadaveric studies (16).

The second theory states that the pain fibres of the MN and the efferent fibres of the shoulder could potentially be sharing a second order neuron. This could be triggering the brain to misinterpret the shoulder as a site of origin of the pain (17). Another theory of unmasking the shoulder pain pathways following activation of these pain fibres at the wrist level could be adopted from the original description (18). Moreover, the individual variations in perception of shoulder pain may be explained by combination of all these mechanisms or in different combinations and/or in different intensity. Our study conforms to above theories and observations by showing similar recovery rates of shoulder symptoms on par with hand symptoms in patients with carpal tunnel syndrome.

A systematic review conducted in 2008 revealed that complete or marked improvement occurs in 70-90% of patients by one year following treatment for CTS (19). Recovery rate of hand symptoms by day-21 in our study was 85-89%.

Recovery rates of hand symptoms between the two groups showed no significant difference. Therefore, despite having an additional shoulder symptom both groups appeared identical. Furthermore, shoulder symptoms recovered at the same rate. There was no statistically significant difference in their hand-symptom recovery rates and shoulder-symptom recovery rates. These results have two implications to patients with CTS. First, patients with typical shoulder symptoms of CTS have a good recovery following MN decompression at the carpus. Secondly, recovery from hand symptoms and shoulder symptoms of CTS may take up to a few weeks for complete resolution. Which means that there is no immediate complete resolution in many patients. This is explained by the nerve symptoms that are thought to have the ability to register themselves along the pathway (20).

Despite those individual variations, clinicians should offer early decompression and assess potential resolution of shoulder symptoms for a few weeks before subjecting them to advanced investigations. It is a therapeutic option for shoulder symptoms in patients who have already been diagnosed with CTS.

## Limitations

The recovery rates are only followed up to three weeks post decompression.

## Conclusion and recommendations

In our study population, shoulder symptoms of CTS were mainly an aching type, moderately severe pain which started either before or after hand symptoms. The study data showed similar recovery rates for hand symptoms and shoulder symptoms of CTS within the group and between the two groups. The evidence of recovery patterns confirm that these two groups are identical. The occurrence of shoulder pain could be related to interconnection between first order neurons or beyond. The exact mechanism of shoulder symptoms of CTS needs to be further investigated. We recommend considering early decompression of CTS before embarking on additional expensive tests for typical CTS related shoulder symptoms.

## Data Availability

All data produced in the present study are available upon reasonable request to the authors

## Notes

### Competing Interest Statement

The authors have declared no competing interest.

### Funding Statement

This study did not receive any funding

### Author Declarations

Ethical Review Committee of the Teaching Hospital Kurunegala, Sri Lanka

